# NHAPL analysis of glycoRNA reveals sialic acid-containing glycosylated mRNA 3′UTRs and enables sensitive SLE diagnostics

**DOI:** 10.64898/2026.02.05.26345357

**Authors:** Jie Gui, Meng Zhang, Ziwei Kan, Xiaojuan He, Meipei Gao, Jian Han, Qiongfang Wang, Shengyao Zhang, Junyi Hu, Wenyi Qin, Zi Bi, Boyue Huang, Zhongjun Wu, Jianhua Ran

**Affiliations:** Department of Anatomy, and Laboratory of Neuroscience and Tissue Engineering, Basic Medical College, Chongqing Medical University, Chongqing, China; Department of Hepatobiliary and Pancreatic Surgery, The First Affiliated Hospital of Chongqing Medical University, Chongqing, China; State Key Laboratory of Natural Medicines, School of Basic Medicine and Clinical Pharmacy, China Pharmaceutical University, Nanjing, China; School of Basic Medical Sciences, Guangzhou Medical University, Guangzhou, 511436, China; Cancer Science Institute of Singapore, National University of Singapore, 117599, Singapore; Department of Integrated Traditional Chinese and Western Medicine, The First Affiliated Hospital of Chongqing Medical University, Chongqing, China; University-Town Hospital of Chongqing Medical University, Chongqing, China; Western Institute of Digital-Intelligent Medicine, Chongqing, China; Key Laboratory of Major Brain Disease and Aging Research (Ministry of Education), Chongqing Medical University, Chongqing, China

**Keywords:** GlycoRNA, mRNA N-glycosylation, 3′ untranslated region (3′UTR), Systemic lupus erythematosus (SLE), Cell surface RNA, NHAPL

## Abstract

GlycoRNA, newly identified RNA molecules bearing glycan modifications on cell membranes, is implicated in cell communication and immune regulation. However, current technological limitations impede a thorough elucidation of their biological roles and clinical significance. Here, we developed <u>N</u>ucleotides <u>H</u>ybridization and <u>A</u>ptamer-based <u>P</u>roximity <u>L</u>igation (NHAPL), a homogeneous assay enabling sensitive and quantitative glycoRNA analysis from 160pg total cell RNA and 1µl serum. NHAPL integrates dual recognition by a sialic acid aptamer and RNA binding probe, followed by ligation and qPCR amplification. We further established multiplexed NHAPL for simultaneous detection of multiple glycoRNA. Using NHAPL, we uncover for the first time that protein-coding mRNAs, specifically 3′ untranslated region (3′UTR) fragments of FNDC3B and CTSS, undergo sialic acid-containing N-glycosylation on the cell surface. These glycoRNAs functionally promote monocyte adhesion to endothelial cells and hepatoma cell migration, revealing a direct role in cell–cell interactions and cancer-related phenotypes. Applying multiplexed NHAPL to human serum, we identify glycoRNA signatures highly specific to systemic lupus erythematosus (SLE). In particular, glycoY5 and glycoU1 achieve near-complete discrimination between patients and healthy controls (area under the curve (AUC) = 1.00 and 0.9977), whereas conventional total RNA analysis fails to capture these differences, highlighting RNA glycosylation modification as a distinct regulatory layer. Its simplicity and flexibility make it well suited for clinical glycoRNA profiling and biomarker discovery. Overall, NHAPL represents a robust and versatile platform for advancing glycoRNA research and diagnostic development.

## 2 Introduction

Recent studies have revealed that cells can glycosylate small noncoding RNAs—including tRNAs, Y RNAs, snRNAs, snoRNAs, rRNAs, and miRNAs—enriched in sialic acid and fucose. This discovery has established a new class of glycoconjugates, termed glycoRNA, alongside proteoglycans and glycosphingolipids(Flynn et al., 2021; J. Li et al., 2023). N-glycosylation has been mapped to 3-(3-amino-3-carboxypropyl) uridine (acp^3^U) (Xie et al., 2024), and evidence suggests that glycoRNA are glycosylated in the endoplasmic reticulum, processed through the ER–Golgi pathway(Z. Ren et al., 2023), and displayed on the cell surface where they engage with proteins such as Siglecs(Flynn et al., 2021; Liu et al., 2024), P-selectin(N. Zhang et al., 2024), and other RNA-binding receptors(Perr et al., 2025).

Despite their emerging significance(Graziano et al., 2025; Perr et al., 2025; N. Zhang et al., 2024), progress in glycoRNA biology has been constrained by the lack of effective analytical tools. Current methods fall into two categories: (i) in vitro labeling and enrichment approaches— including metabolic chemical reporters (MCRs)(Flynn et al., 2021), chemoenzymatic strategies(J. Li et al., 2023), and RNA-adapted periodate oxidation and ligation (rPAL)(Xie et al., 2024), which enable glycoRNA purification for gel blotting(Flynn et al., 2021; Sharma, Jiao, Yang, Kwan, & Kiledjian, 2025), next-generation sequencing(J. Li et al., 2023), or glycan analysis via mass spectrometry(Flynn et al., 2021); and (ii) in situ visualization approaches that detect glycoRNA in single cells(Liu et al., 2024; Ma et al., 2023) or extracellular vesicles(T. Ren et al., 2025). While powerful, these methods largely rely on purified samples, membrane-dependent imaging, or separate analyses of RNA and glycan components, and therefore they cannot provide glycoRNA information when the membrane does not need to remain intact (i.e., membrane-independent) or in cell-free contexts, and they are not well suited for diagnostics.

To address these challenges, we developed <u>N</u>ucleotides <u>H</u>ybridization and <u>A</u>ptamer-based <u>P</u>roximity <u>L</u>igation (NHAPL), a homogeneous assay that integrates a previously reported sialic acid–specific aptamer (Yue et al., 2021), which selectively binds sialylated N-glycans with reported dissociation constants of 87.34 ± 15.19 nM (Yue et al., 2021) and 91 nM (Ma et al., 2023), with a complementary DNA probe for dual recognition of glycoRNA. This dual-binding event is transduced via connector-mediated proximity ligation(Fredriksson et al., 2002), followed by qPCR amplification (Fig. 1a, b). Systematic optimization of the assay parameters confers high specificity and sensitivity.

**Fig. 1.**
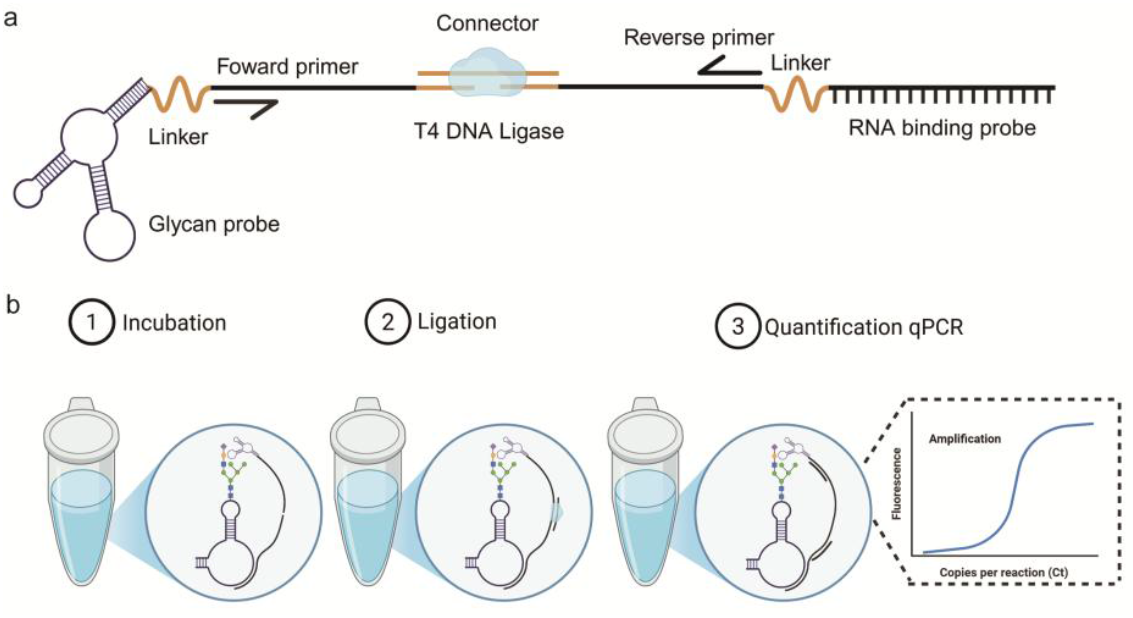
Principle of NHAPL. **a**, Architecture and domain composition of the glycan probe, RNA binding probe, and other components. **b**, Principle of NHAPL highlighting homogenous glycoRNA detection and signal transduction.

After validating NHAPL in total RNA from cultured cells, we extended the method to a multiplexed format for simultaneous detection of multiple glycoRNA. Using this platform, we further investigated mRNA N-glycosylation and identified sialylated N-linked glycans in the 3′UTR fragments of *FNDC3B* and *CTSS* (termed glyco*FNDC3B* and glyco*CTSS*) both in THP-1 and Hep3B cells. This finding expands previous knowledge that glycoRNA are limited to small noncoding RNAs(Flynn et al., 2021). To probe their function, we inhibited glyco*FNDC3B* and glyco*CTSS* by co-culturing THP-1 cells with antisense oligonucleotides. This treatment significantly reduced adhesion to HUVECs, consistent with earlier observations(Huang et al., 2020).Furthermore, pretreating live cells with RNase or NGI-1 to remove cell surface RNA and glycan moiety significantly decreased glyco*FNDC3B* and glyco*CTSS* signals, indicating that these glycoRNA are surface-displayed. Given that *FNDC3B* promotes cell migration and metastasis in hepatocellular carcinoma(C. H. Lin et al., 2016), we examined the role of glyco*FNDC3B* in cancer cell behavior. Strikingly, its inhibition markedly decreased migration, implicating glyco*FNDC3B* possess a function in tumor progression. Finally, we demonstrated that glycoRNA are stably present in serum, with highly consistent expression levels among healthy individuals (Pearson’s R >0.8). Importantly, glycoY5 and glycoU1 exhibited excellent diagnostic performance in SLE, with AUROC values of 1 and 0.9977 respectively, highlighting glycoRNA as a promising new class of disease biomarkers.

## 3 Methods

### 3.1 Materials

Sodium chloride, magnesium chloride, 2-mercaptoethanol, chloroform, NGI-1, kifunensine, swainsonine and Tween-20 were purchased from Sigma-Aldrich. Ac4ManNAz, dibenzocyclooctyne-PEG4-biotin (DBCO-biotin), penicillin, streptomycin, high sensitive Plus ECL luminescence reagent and fetal bovine serum were purchased from Sangon Biotech (Shanghai) Co., Ltd. T4 DNA ligase, ATP, DTT, proteinase K, PNGase-F were purchased from New England BioLabs (NEB). RNase A and RNase T1 were purchased from Thermo Fisher Scientific. RNAiso Plus was purchased from Takara. Formamide, isopropanol and ethanol were purchased from Sichuan East Chemical Industry. Phorbol-12-myristate-13-acetate (PMA), lipopolysaccharide (LPS), blocking buffer, streptavidin and 2x RNA loading buffer were purchased from Beyotime. Hifair® III 1st Strand cDNA Synthesis SuperMix for qPCR(gDNA digester plus) were purchased from Yeasen Biotechnology (Shanghai) Co., Ltd. The 0.45-µm nitrocellulose (NC) membrane was purchased from Cytiva Life Sciences. QPCR reaction Mix and 50 bp DNA Ladder was purchased from Beijing Tsingke Biotech Co., Ltd.

The Neu5Ac-specific aptamer was adapted from previously reported sequences, exhibiting affinities of 87.34 ± 15.19 nM(Yue et al., 2021) and 91 nM(Ma et al., 2023). RNA-binding regions for glycoU1, U3, U35, U8, and Y5 were adopted from Ref.(Ma et al., 2023), while those for FNDC3B and CTSS were adapted from Ref.(Huang et al., 2020). All the oligonucleotide sequences were purchased from Sangon Biotech (Shanghai) Co., Ltd. and were purified by high-performance liquid chromatography or polyacrylamide gel electrophoresis and confirmed by mass spectrometry (Supplementary Table S1, S2 and S3).

### 3.2 Cell culture

The THP-1 cell line and Hep3B cell line were purchased from ZQXZBIO, Shanghai, China. HUVEC-SV40 was purchased from sunncell, Wuhan, China, authenticated by STR profiling.

Hep3B (ZQ0024) and HUVEC-SV40 (SNL-503) cell lines were cultured in DMEM supplemented with 10% fetal bovine serum (FBS; E600001), 100 U ml^−1^ penicillin and 100 U ml^‒ 1^ streptomycin (E607011). The human monocyte cell line THP-1 (ZQ0086) was cultured in RPMI-1640 medium supplemented with 2.5 mM glutamine, 1× MEM NEAA and 10% heat-inactivated FBS. To differentiate THP-1 cells into macrophage-like cells, cells were treated with 250 nM PMA (S1819) in 10% FBS culture medium for 24 h and treated with 5% FBS RPMI-1640 medium for another 48h. To activate M0 macrophages, macrophages were incubated with 12.5 µg ml^−1^ LPS (S1732) in 5% FBS RPMI-1640 medium for 24h. For RNA blot, cells were labeled with 100nM Ac4ManNAz in medium for at least 24h before RNA extraction. To prepare M0 RNA samples for NHAPL validation, M0 macrophages were treated with 0.02 µg µl^−1^ RNase A (EN0531), 1 U µl^−1^ RNase T1 (EN0541) and 500U ml^-1^ PNGase-F (P0704) in PBS at 37 °C for 30 min. After incubation, the cells were washed in PBS three times, followed by RNA extraction.

To remove the glycan moiety, both pharmacological and enzymatic methods were adopted according to previous report(Ma et al., 2023). Live cells were incubated with (1) 8 µM NGI-1, which is a specific small-molecule inhibitor of oligosaccharyltransferase related to glycoRNA generation(Lopez-Sambrooks et al., 2016); (2) 2 µM kifunensine, which can inhibit N-glycan processing(Elbein, Tropea, Mitchell, & Kaushal, 1990); or (3) 40 µM swainsonine, which can perturb N-glycan processing(Tulsiani, Harris, & Touster, 1982) for 24 h before the RNA extraction experiment. In the enzymatic method, to digest glycans on the cell surface of live cells, cells were incubated with 100 U ml^−1^ PNGase-F at 37 °C for 30 min.

To remove the RNA moiety, live cells were incubated with 0.02 µg µl^−1^ RNase A (EN0531), 1 U µl^−1^ RNase T1 (EN0541) for 20min, followed by washing thoroughly in PBS for three times before RNA extraction.

### 3.3 Total RNA extraction

Total RNA of cells or serum was extracted by RNAiso Plus (9108) and SanPrep Column microRNA Extraction Kit (B518811-0050) according to the manufacturer’s instructions, and quantified by Nanodrop 2000 Spectrophotometer (Thermo Scientific). Notably, 200µl of each serum sample were added to 2ml RNAiso Plus (9108). Since U6 and 5S rRNA were degraded in serum samples, and there is no current consensus on the use of house-keeping RNA for RT-qPCR analysis, the expression levels of target RNA were directly normalized to serum input. Impure total RNA samples might inhibit enzymatic ligation of T4 DNA ligase, leading to the failure of NHAPL assays.

### 3.4 RNA blot of Ac4ManNAz-labeled RNA

GlycoRNA in THP-1 cells, M0 macrophages, and M1 macrophages were examined by metabolic labeling with Ac4ManNAz followed by RNA blotting, as previously reported(Flynn et al., 2021; Li, Zhang, Pantoja, Wang, & Lu, 2024). Briefly, total RNA was extracted after labeling, treated with proteinase K to remove protein contaminants, and conjugated with DBCO-biotin via bioorthogonal click chemistry. The RNA was then purified by chloroform extraction and SanPrep Column microRNA Extraction Kit (B518811-0050), and subsequently analyzed by denaturing gel electrophoresis and RNA blotting.

#### Metabolic labeling

For the RNA blotting experiments, metabolic labeling by Ac4ManNAz was performed by culturing cells with 100 µM Ac4ManNAz in cell culture medium for 24-36h before RNA extraction.

#### RNA extraction and purification

RNA extraction and purification were performed according to a previously reported method(Flynn et al., 2021). Cells after Ac4ManNAz labeling were first treated with 1 ml of RNAiso Plus (9108) and incubated at room temperature to denature non-covalent interactions. Phase separation was then performed by adding 0.2 ml of chloroform, followed by vortexing and incubating for 5 min on ice, then centrifuging at 12,000g at 4 °C for 15 min. The RNA in the aqueous phase was carefully transferred to a fresh tube and purified using a SanPrep Column microRNA Extraction Kit (B518811-0050). To avoid protein contamination, the obtained RNA was subjected to protein digestion by adding 1 µg of proteinase K to 25 µg of purified RNA in 30 mM Tris-HCl (pH 8.0) and incubated at 37 °C for 45 min. To remove the proteinase K, the RNA was purified again with a SanPrep Column microRNA Extraction Kit (B518811-0050) and stored at –80 °C for future use.

#### Copper-free click conjugation to Ac4ManNAz-labeled glycoRNA

Ac4ManNAz-labeled RNA (10 µl) was mixed with 10 µl RNA denature buffer (95% formamide, 18 mM EDTA and 0.025% SDS) and 1.5 µl of 10 mM DBCO-biotin. Conjugation was performed at 37 °C for 30 min in a PCR incubator. Biotin-labeled RNA was then purified with a SanPrep Column microRNA Extraction Kit (B518811-0050).

#### RNA gel electrophoresis, blotting and imaging

The gel electrophoresis and blotting analysis of biotin-labeled RNA was performed according to previously reported methods(Flynn et al., 2021; Li et al., 2024). Purified biotin-labeled RNA was mixed with 2x RNA loading buffer (R0215) and incubated at 55 °C for 10 min and snapped cooled on ice. Then, 2 µg of RNA was loaded into 1% agarose-formaldehyde denaturing gels, electrophoresed at 110 V for 30min on ice and visualized after electrophoresis. The RNA sample was transferred to a 0.45-µm NC membrane using the same method as the bottom-up transfer for northern blotting at room temperature for 12-24h. Afterward, the RNA sample was cross-linked to the NC membrane with UV cross-linker for 30 min. The NC membrane was then blocked with blocking buffer (D3308B) at room temperature for 1h and stained with streptavidin (A0303) in blocking buffer (1:10,000 dilution) at room temperature for 1h. The NC membrane was then washed with 1× PBST three times. The NC membrane was then imaged with high sensitive Plus ECL luminescence reagent (C520045). The gel and blot images were measured using ImageJ and prism 8 software

### 3.5 Single NHAPL assays

Total RNA samples were diluted with incubation buffer(Guo et al., 2025) (50 mM Tris-HCl, 5 mM KCl, 100 mM NaCl and 1 mM MgCl2, pH 7.4 @ 25°C) to corresponding concentration. 1ul RNA samples were added to 2ul incubation mix (150pM glycan binding probe, 75pM RNA binding probe, 50 mM Tris-HCl, 5 mM KCl, 100 mM NaCl and 1 mM MgCl_2_, pH 7.4) and incubated for 1h at room temperature. Then, 47ul ligation mix (0.5 weiss units T4 DNA ligase, 400nM connector, 50 mM Tris-HCl, 10 mM MgCl_2_, 1 mM ATP, 10 mM DTT, 0.05% BSA, pH 7.4 @ 25°C) were added and incubated for 5min at room temperature. After ligation, T4 DNA ligase was heat inactive at 65°C for 10min. Then, a 3μl aliquot of the reaction product was added to 17 μl of qPCR reaction Mix (DLQ104) with 500nM primers. SYBR Green-based qPCR was performed on a Archimed real-time PCR detection system (95 °C for 3 min, 95 °C for 15 s, 60 °C for 1 min, 40 cycles).

For RNA concentration assay, 100pM glycan binding probe, 100pM RNA, 50nM connector and 0.1 weiss units T4 DNA ligase was used. For glycan probe, RNA binding probe, connector and T4 DNA ligase concentration selection experiments, all the remaining conditions are the same as RNA concentration assay except corresponding components to be optimized.

To evaluate the performance of NHAPL, total RNA from M0 macrophages was mixed with exponentially increasing amounts of THP-1 RNA (1 pg, 2 pg, 4 pg, 8 pg, …) while maintaining a constant total RNA input of 160 pg. The mixed RNA samples were then subjected to NHAPL analysis.

### 3.6 Multiplexed NHAPL assays

The protocol was similar to single NHAPL assays with minor modifications. Briefly, 1ul RNA samples were added to 2ul incubation mix (150pM glycan binding probe, 75pM multiplexed RNA binding probe set, 50 mM Tris-HCl, 5 mM KCl, 100 mM NaCl and 1 mM MgCl_2_, pH 7.4 @ 25°C) and incubated 1h at room temperature. To acquire a better Ct readout, extending the incubation time is recommended. Then, 47ul ligation mix (0.5 weiss units T4 DNA ligase, 400nM connector, 50 mM Tris-HCl, 10 mM MgCl2, 1 mM ATP, 10 mM DTT, 0.05% BSA, pH 7.4 @ 25°C) were added and incubated for 5min at room temperature. After ligation, T4 DNA ligase was heat inactive at 65°C for 10min. A 3μl aliquot of the reaction product was added to 17 μl of qPCR reaction mix (DLQ104) with 500nM universal primer and specific primer respectively. SYBR Green-based qPCR was performed on a Archimed real-time PCR detection system (95 °C for 3 min, 95 °C for 15 s, 60 °C for 1 min, 40 cycles).

### 3.7 Clinical sample analysis

This study involved the use of human serum samples. The study protocol was reviewed and approved by the Ethics Committee of the First Affiliated Hospital of Chongqing Medical University (Chongqing, China). The review was conducted through a convened ethics committee meeting, and ethical approval was granted (Approval No.: 2024-029-01). All procedures involving human participants were conducted in accordance with the ethical standards of the institutional research committee and with the Declaration of Helsinki. Written informed consent was obtained from all participants prior to sample collection.

Whole blood samples of SLE patients, IIM, viral hepatitis or healthy donors were obtained from the First Affiliated Hospital of Chongqing Medical University (2024-029-01) and University-Town Hospital, Chongqing Medical University (LL-202440), respectively. Notably, all the 20 SLE serum samples were obtained from hospitalized patients with a confirmed diagnosis of SLE (Table S4).

The blood samples were first centrifuged at 1,550g for 15 min to obtain cell-free serum and stored at -80°C before use.

Briefly, 1 μl serum was incubated with 1ul 2x preincubation buffer (100 mM Tris-HCl, 10 mM KCl, 200 mM NaCl and 2 mM MgCl_2_, 4% BSA, 32ug ml^-1^ poly(A), 0.1% Tween-20, pH 7.4 @ 25°C) for 20min at room temperature. Then, 1μl preincubated sample was analyzed by multiplexed NHAPL as described above. For serum analysis, each sample was tested twice using multiplexed NHAPL.

For serum heat treatment, after incubation at 80°C for 10min, 1μl supernatant was subjected to multiplexed NHAPL analysis as described above. In this experiment, serum samples from one healthy volunteer and two patients with systemic lupus erythematosus (SLE) were analyzed.

### 3.8 Serum total RNA analysis

Total serum RNA was isolated from 200ul serum using a SanPrep Column microRNA Extraction Kit (B518811-0050) according to the manufacturer’s instructions. Then, reverse transcriptase reactions were conducted using Hifair® III 1st Strand cDNA Synthesis SuperMix for qPCR (gDNA digester plus) Kit according to the manufacturer’s instructions, followed by qPCR analysis. All the primers used were list in Table S3.

### 3.9 THP-1 attachment assay

THP-1 cells were stained with Calcein AM (2.5 μM, Beyotime, C2012) for 30 min at 37 °C for live-cell fluorescent tracking. Meanwhile, antisense probes were denatured at 80 °C for 5 min, and snap cooled on ice. After incubation, THP-1 cells were washed and resuspended in serum-free RPMI media at 10^6^ cells mL^-1^ and evenly segregated for hybridization with 10 nM antisense probes for 30min at 37 °C.

After incubation, THP-1 cells were washed thoroughly in PBS for three times to remove unbound antisense oligos and were seeded onto a confluent layer of HUVECs in 96-well plates. THP-1 cells were incubated for 30 min to allow for attachment. Unattached cells were removed with PBS washes. Attachment of the fluorescently labeled THP-1 cells was quantified using the Spark^®^ Multimode Microplate Reader according the manufacture’s instruction. Statistical summary and tests were performed in Prism 8. P values were generated by Kruskal-Wallis nonparametric test for multiple comparisons against the control groups with Bonferroni adjustments.

### 3.10 Wound healing assay

Antisense probes for *FNDC3B* and *CTSS* were denatured at 80 °C for 5 min, and snap cooled on ice. Live Hep3B cells were cultured to 80% confluent, subsequently co-cultured with 10nM antisense probes (a scrambled antisense oligo as negative control), respectively. The wound healing assay was performed using a 200 μl pipette tip to scratch the Hep3B cell layer, and then washed 3 times with PBS to remove cell fragments. The wound healing rate was recorded after 0, 12, 24, 36 and 48 h. The wound distances were measured using ImageJ and Prism 8 software. All experiments were independently repeated three times.

### 3.11 Statistical analysis

All data are represented as mean ± SD. Statistical analyses were performed using SPSS (version 17), Origin (version 2021) and GraphPad Prism (version 8.0) software. Data were analyzed under the Student’s t-test (2-tailed), ANOVA or Pearson’s correlation coefficient (R) methods. *P*-value of less than 0.05 was considered as statistically significant. ns (not statistically significant), *(P < 0.05), ** (P < 0.01), *** (P < 0.001), **** (P < 0.0001). The schematics in Figs. 1, 2a, 2j, 3a, 4a, 4b, 5a, 5f, 6a, Extended fig 1a, 2a, 4a and 5a were created with BioRender.com. A BioRender academic license for using these artworks for publication is in place.

**Fig. 2.**
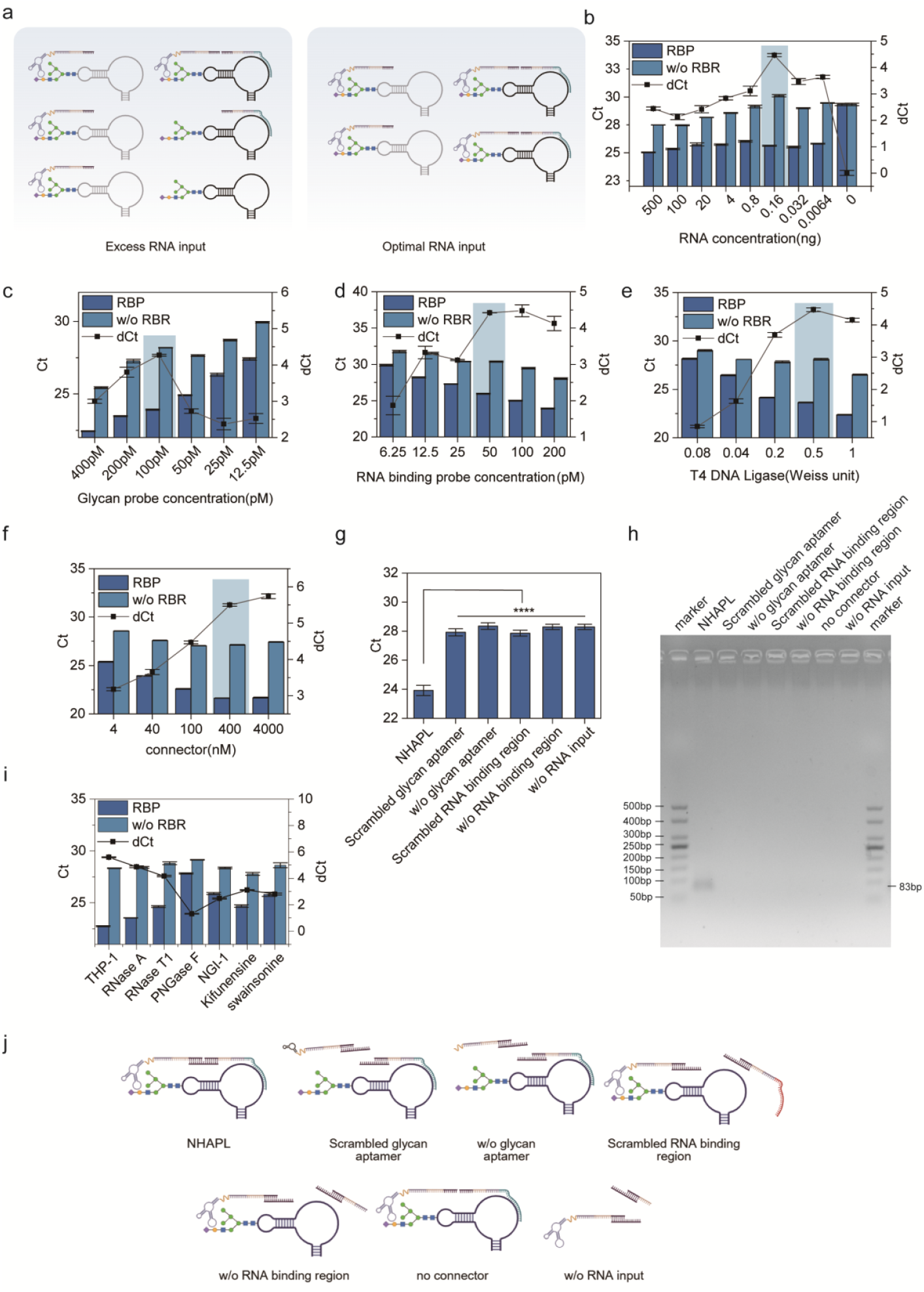
Optimization of NHAPL for homogeneous glycoRNA detection. **a**, Schematic showing the effect of total RNA input on probe binding efficiency in NHAPL. **b**, NHAPL Ct and dCt values across varying RNA inputs; The left y-axis is the NHAPL signal Ct. The right y-axis is the NHAPL signal dCt calculated by subtracting the Ct value of the RNA binding probes for U1 (RBP) from that of RNA binding probes without RNA binding region (negative control, w/o RBR). **c–f**, Optimization of assay conditions by varying the concentrations of glycan probe, RNA binding probe, T4 DNA ligase, and connector. **g, h, j** Ct values, gel electrophoresis of PCR products after 25 amplification cycles and schematic illustration of validation of control groups lacking individual components. NHAPL signals were undetectable in the absence of connector. **i**, NHAPL analysis of glycoRNA abundance modulated through both enzymatic and pharmacological treatments. Error bars are not visible for most data points due to minimal variance. Data shown as mean ± s.d. (n = 3). w/o, without. ns (not statistically significant), *(P < 0.05), ** (P < 0.01), *** (P < 0.001), **** (P < 0.0001).

## 4 Results

### 4.1 Development of NHAPL for homogenous detection of glycoRNA

To enable homogenously quantitative detection of glycoRNA, we developed <u>N</u>ucleotides <u>H</u>ybridization and <u>A</u>ptamer-based <u>P</u>roximity <u>L</u>igation (NHAPL), an adaptation of the proximity ligation assay (PLA)(Fredriksson et al., 2002; Gullberg et al., 2004) for in vitro analysis of glycoRNA. NHAPL integrates four key components (Fig. 1a): (1) a glycan probe carrying a Neu5Ac-specific aptamer, adapted from previously reported sequences (Ma et al., 2023; Yue et al., 2021), which selectively binds sialylated N-glycans with a reported dissociation constant of 87.34 ± 15.19 nM, enabling recognition of sialylated glycoRNA. (Flynn et al., 2021), a linker to avoid steric hindrance during recognition, a primer binding site for subsequent qPCR analysis and a connector hybridization site. (2) an RNA binding probe with a RNA hybridization region complementary to the target glycoRNA sequence, a linker to avoid steric hindrance during hybridization, a primer binding site for qPCR analysis and a connector hybridization site. (3) a connector oligonucleotide bridging both probes by hybridizing to their free connector hybridization site. (4) a primer pair for subsequent qPCR amplification. Dual recognition of both glycan probe and RNA binding probe to the same target glycoRNA brings their free ends into proximity to allow connector hybridization and enzymatic ligation. The ligated product is then amplified by qPCR, serving as a quantitative readout (Fig. 1b). This process generates ligated products proportional to the initial concentration of the target glycoRNA (signal ligations), along with a smaller fraction of products (background ligations) arising from the infrequent joining of both glycan probe and RNA binding probe in the absence of specific glycoRNA(Gustafsdottir et al., 2005) (Extended Fig. 1a). Although qPCR cannot distinguish between signal and background ligations, under optimized conditions the signal products predominate, enabling ultrasensitive and indirect detection of the target glycoRNA. Signal amplification in NHAPL relies on the proximity effect, where simultaneous binding of two probes to the same glycoRNA markedly increases their local effective concentration compared with unbound free probes, thereby enhancing ligation efficiency(Fredriksson et al., 2002; Gullberg et al., 2004; Gustafsdottir et al., 2005).

NHAPL was first validated with small nuclear RNA U1 modified with sialic acid-containing N-linked glycans (glycoU1) as a representative target in a fully homogeneous format without washing, separating or imaging procedures. Since the abundance of glycoRNA in total cellular RNA was unknown, we first investigated the effect of total RNA input on the detection performance. RNA input optimization experiments revealed that excessive RNA input reduced specificity due to glycan probe competition by massive glycans from non-target glycoRNA (Fig. 2a, b). A concentration assay identified 160 picogram (pg) total RNA as optimal for signal-to-noise performance (Fig. 2b). When the total RNA input was excessive, glycans from non-target glycoRNA competed with those on the target molecules for binding to the glycan probe, thereby reducing the effective binding efficiency. In contrast, decreasing the total RNA input minimized this competition, allowing the glycan probe to bind more efficiently to the target glycoRNA and achieve higher detection sensitivity. However, reducing the total RNA input also decreased the overall abundance of glycoRNA in the sample, thereby limiting the number of target glycoRNA available for hybridization with the RNA binding probe and consequently reducing its detection efficiency (Fig. 2a). Taken together, decreasing the total RNA input did not quantitatively increase the glycoRNA-dependent signal (Ct value) (Fig. 2b). However, as the total RNA input decreased, the Ct value of the negative control (RNA binding probe without RNA binding region, w/o RBR) increased almost linearly and reached a plateau when the RNA input was reduced to less than 800 pg (Fig. 2b). Consistent results were obtained when maintaining a fixed RNA input of 160 pg while supplementing the remaining nucleic acid content with salmon sperm DNA (Extended Fig. 1b). As PLA relies on an excess of connector oligonucleotides to rapidly hybridize with unbound probes to minimize nonspecific hybridization and suppress the generation of glycoRNA-independent ligation substrates(Fredriksson et al., 2002; Gullberg et al., 2004), excessive total RNA can hinder connector hybridization to free probes and increase background signals, while an optimal RNA input avoids this effect (Fig. 2b, Extended Fig. 1a, b).

Given that sialylation on RNA varies across cell types (12–16 residues in HEK293T vs. 4– 9 in HeLa for glycoY5)(Liu et al., 2024), the two probe ratios were further optimized. Unlike conventional protein PLA requiring equimolar probes, glycoRNA detection required excess glycan probe for universal application. The best performance was achieved by incubating RNA samples with 100 pM glycan probes and 50 pM RNA binding probes, along with 400 nM connector and 0.5 Weiss unit T4 DNA ligase (refer to the Methods section for details) (Fig. 2c– f). Excess amounts of both probes or ligase markedly increase nonspecific ligation events, thereby reducing the dynamic detection range.

Previous studies have suggested the presence of glycoprotein contamination in total RNA samples(Kim et al., 2024). Therefore, we examined whether potential glycoproteins in total RNA could affect NHAPL analysis (Extended Fig. 2 a-d). As shown in Extended Fig. 2d, total RNA was treated with proteinase K and subsequently repurified, and the NHAPL signals of the protein-depleted RNA showed no significant difference compared with the untreated control group. Since NHAPL relies on the simultaneous binding of two probes to the same glycoRNA, which markedly increases their local effective concentration relative to unbound free probes (including those binding to free glycans, as shown in Extended Fig. 2c), ligation efficiency is enhanced and only one ligation product is generated per glycoRNA because each glycoRNA only complementary to one RNA binding probes. Thus, glycoRNA-binding protein indeed did not affect NHAPL analysis (Extended Fig. 2 a). Proteinase K treatment data ruled out the existence of glycol–protein–RNA complexes (that is, unglycosylated RNAs associated with glycoproteins) (Extended Fig. 2b, d). If such complexes existed, the close proximity between the glycans on glycoproteins and the RNA could potentially generate NHAPL signals. However, proteinase K digestion would disrupt these complexes, thereby eliminating any NHAPL signals arising from this source.

Specificity controls confirmed that omission of any essential component—glycoRNA, probe, or connector—led to a marked reduction or complete loss of detectable ligation products (Fig. 2g, j). Gel electrophoresis (Fig. 2h) verified the expected amplicon size, confirming that qPCR readouts accurately reflected the abundance of ligation products. To further validate the performance and accuracy of NHAPL for glycoRNA detection, glycoRNA abundance was modulated through both enzymatic and pharmacological treatments. Removal of the glycan moiety by treating live cells with PNGase F, a glycosidase that cleaves N-linked glycans (Flynn et al., 2021; Ma et al., 2023), yielded almost undetectable NHAPL signals (fig. 2i), indicating nearly complete deglycosylation of cell surface glycoRNA. Likewise, pretreatment with glycosylation inhibitors (NGI-1, kifunensine, or swainsonine) for 24 h substantially reduced NHAPL signals, consistent with previous reports(Ma et al., 2023). Moreover, to validate NHAPL results, click chemistry and metabolic labeling-based blots were performed and obtained the similar glycoRNA expression trend (Extended Fig. 3). To assess the dependence on the RNA component, cells were incubated with RNase A or RNase T1 at 37 °C for 20 min before analysis. Interestingly, only a slight decrease in NHAPL signal was observed (fig. 2i), in contrast to previous findings(Ma et al., 2023). This mild reduction, also supported by metabolic labeling with Ac4ManNAz followed by biotinylation and RNA blotting(Flynn et al., 2021) (Extended Fig. 3), suggests that glycoRNA possess partial RNase resistance(T. Ren et al., 2025; N. Zhang et al., 2024). Collectively, these results demonstrate that NHAPL provides a robust and specific method for homogeneous glycoRNA detection.

### 4.2 NHAPL exhibited robust analytical sensitivity

Beyond qualitative assessment, we conducted quantitative validation of NHAPL. Two considerations guided this analysis: (i) NHAPL relies on qPCR, yielding logarithmic signals, and (ii) standard glycoRNA controls are unavailable due to difficulties in in vitro synthesis. We found that PMA-induced differentiation of THP-1 cells into M0 macrophages resulted in negligible glycoRNA expression using RNA blot(Li et al., 2024) (Fig. 3a, b), consistent with previous findings(Ma et al., 2023). Thus, M0 macrophages were used to generate negative controls by pretreating live M0 macrophages with RNase A, RNase T1 and PNGase-F, ensuring surface glycoRNA removal(Ma et al., 2023). NHAPL analysis confirmed that signals for glycoU1, U3, U35a, Y5 and miR155(J. Li et al., 2023) were only marginally above background signals, indicating near-complete depletion (Fig. 3c).

**Fig. 3.**
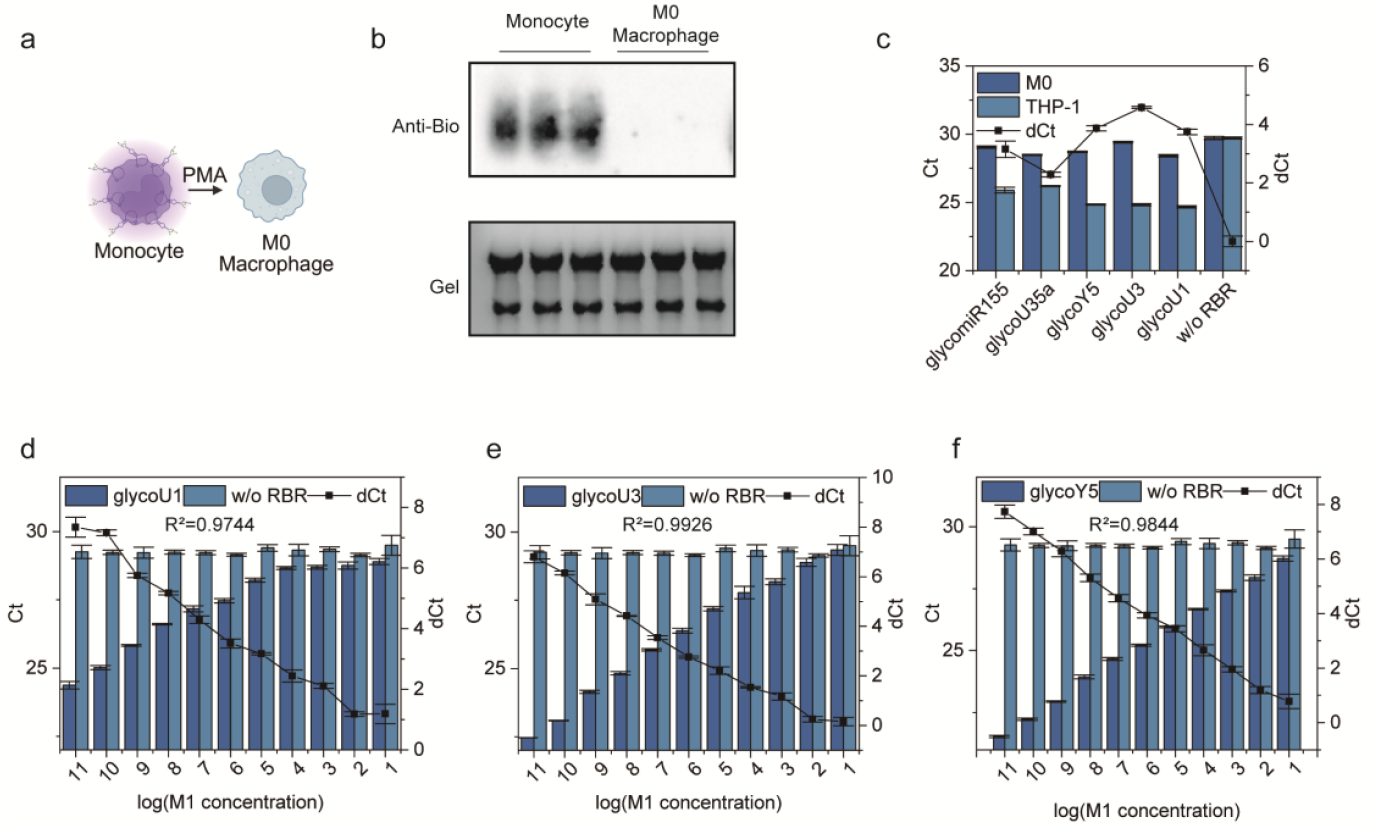
Validation of optimized NHAPL using a glycoRNA gradient model. **a**, Diagram illustrating the differentiation of THP-1 cells. **b**, GlycoRNA expression in monocytes and M0 macrophages was evaluated by metabolic labeling with Ac4ManNAz. Extracted RNAs were subsequently biotinylated via click chemistry and analyzed by agarose gel electrophoresis (lower panel) followed by anti-biotin RNA blotting (upper panel). **c**, Comparison of glycoRNA levels in M0 macrophages pretreated with or without RNase A, RNase T1 and PNGase-F and THP-1 cells by NHAPL. The dCt line is for THP-1 by subtracting the Ct value of the RNA binding probes (for U1, U3, Y5, U35a or miR155) from that of RNA binding probes without RNA binding region (negative control, w/o RBR). **d–f**, Simple linear regression between the NHAPL signals and the THP-1 RNA sample concentration. The left y-axis is the NHAPL signal Ct. The right y-axis is the NHAPL signal dCt calculated by subtracting the Ct value of the RNA binding probes (for U1, U3, Y5, U35a or miR155) from that of RNA binding probes without RNA binding region (negative control, w/o RBR). Considering the qPCR signals were logarithmic in nature, the THP-1 RNA sample concentration was log-transformed and plotted on the x-axis. Error bars are not visible for most data points due to minimal variance. Data shown as mean ± s.d. (n = 3).

To evaluate quantitative performance, M0 macrophage RNA were mixed with exponentially increasing concentrations of THP-1 RNA and analyzed by NHAPL (refer to the Methods section for details). The dCt values correlated strongly with the proportion of THP-1 RNA, yielding R^2^ values of 0.9846 for glycoU1 (95% CI: 0.9695–0.9981), 0.9821 for glycoU3 (95% CI: 0.9821– 0.9989), and 0.9959 for glycoY5 (95% CI: 0.9959–0.9997) (Fig. 3d–f).

Collectively, these results demonstrated that NHAPL enables highly sensitive and specific quantification of glycoRNA, supporting its utility as a reliable analytical tool.

### 4.3 Multiplexed NHAPL enables simultaneous detection of multiple glycoRNA

Our initial experiments confirmed that NHAPL could reliably detect individual glycoRNA from total RNA samples. To extend its utility, we established a multiplexed NHAPL platform capable of analyzing several glycoRNA in a single reaction. As shown in Fig. 4a, proximity probes were designed with free ends containing universal sequence that allowed ligation of 3′- and 5′-free ends through a common connector, while each RNA binding probe carried a unique primer barcode. When the correct glycoRNA was recognized by the glycan probe and RNA binding probe simultaneously, the free ends of both probes were brought into proximity and produced a glycoRNA specific ligation product. These products were subsequently amplified using a universal primer (paired to the glycan probe) and a glycoRNA-specific primer (paired to the glycoRNA unique barcode). Then, the reaction mixture was transferred to different well of the qPCR plate with corresponding primer sets, enabling simultaneous quantitative detection of each target glycoRNA NHAPL signal products via qPCR (Fig. 4b).

**Fig. 4.**
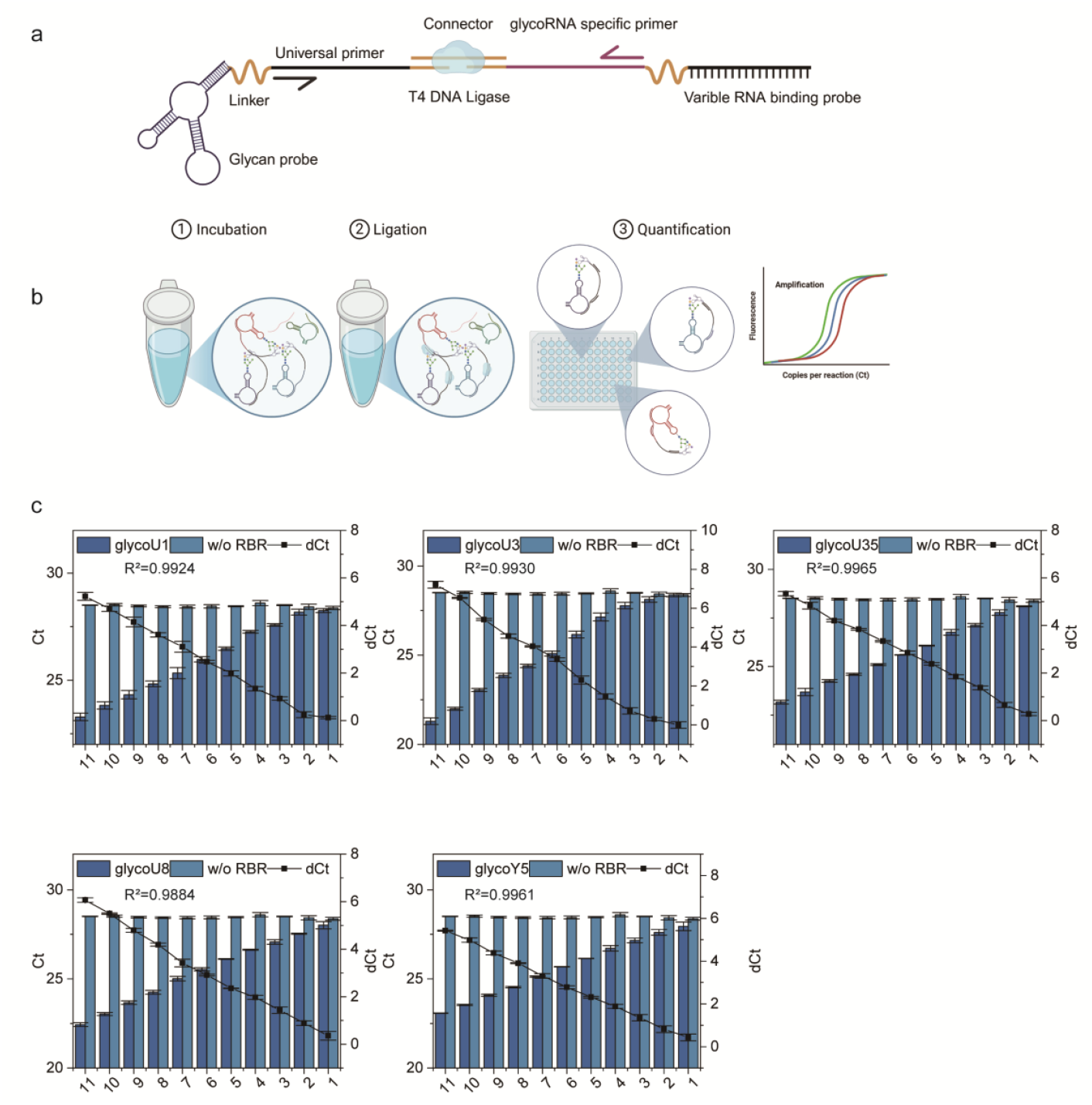
Multiplexed NHAPL analysis of glycoRNA. **a**, Schematic of multiplexed NHAPL, incorporating glycoRNA-specific primer sites in RNA binding probes compared with single NHAPL. **b**, Validation of multiplexed NHAPL using total M0 macrophage RNA mixed with THP-1 RNA fraction. After ligation, reactions were transferred to qPCR plates for real-time fluorescence detection. **c**, Simple linear regression between the multiplexed NHAPL signals and the THP-1 RNA sample concentration. The left y-axis is the multiplexed NHAPL signal Ct. The right y-axis is the multiplexed NHAPL signal dCt calculated by subtracting the Ct value of the RNA binding probes from that of RNA binding probes without RNA binding region (negative control, w/o RBR). Considering the qPCR signals were logarithmic in nature, the THP-1 RNA sample concentration was log-transformed and plotted on the x-axis. Error bars are not visible for most data points due to minimal variance. Data shown as mean ± s.d. (n = 3).

The performance of multiplexed NHAPL was validated by simultaneously detecting glycoU1, U3, U35, U8, Y5 and RNA binding probe without RNA binding region (negative control, w/o RBR) in mixed M0 macrophage RNA samples with varying fractions of THP-1 RNA (refer to the Methods section for details). The assay exhibited strong linearity across all targets (R^2^ = 0.9924 for U1, 0.9884 for U3, 0.9965 for U35a, 0.9930 for U8, and 0.9961 for Y5) (Fig. 4c).

### 4.4 mRNA 3’UTR fractions could be N-glycosylation modified

A variety of small non-coding RNAs, including tRNAs, Y RNAs, snRNAs, and snoRNAs, have been shown to carry sialylated N-linked glycans (glycoRNA) and localize on the plasma membrane(Flynn et al., 2021). Interestingly, previous studies also reported that 3′UTR fragments from coding RNAs, such as those derived from *FNDC3B* and *CTSS*, associate with the cell surface and modulate monocyte adhesion to endothelial cells(Huang et al., 2020). Based on this, we applied NHAPL to examine whether these *FNDC3B* and *CTSS* 3′UTR fragments are glycosylated. Five RNA binding probes complementary to *FNDC3B* or *CTSS* 3′UTR fragments were designed based on previous report(Huang et al., 2020), respectively. Both THP-1 and Hep3B cellular total RNA were analyzed by NHAPL incorporating pretreatment of enzymatic digestion (Fig. 5a). As shown in (Fig. 5b-e), all *FNDC3B* and *CTSS* probes yielded stable glycoRNA signals compared to negative NHAPL control (RNA binding probe without RNA binding region, w/o RBR). Moreover, these glycosylated 3’UTR fractions had the same expression trend both in THP-1 and Hep3B cells, implying a specific glycosylation preference (Fig. 5b–e). When pretreating live cells with PNGase F to remove cell surface glycans (Fig. 5a), the NHAPL signals reduced significantly for all glyco3’UTR fragments (Fig. 5b –e). Notably, pretreating live cells with RNase showed no effect on some of the glyco3’UTR fragments, indicating a RNase resistance capability of these glyco3’UTR fragments, in agreement with (Fig. 2i) and previous report (T. Ren et al., 2025; N. Zhang et al., 2024). The above data indicated that nearly all these glyco3’UTR fragments are sensitive to enzymatic digestion for both glycans and RNA in live cell membrane, demonstrating that these glyco3’UTR fragments are located on cell membrane.

**Fig. 5.**
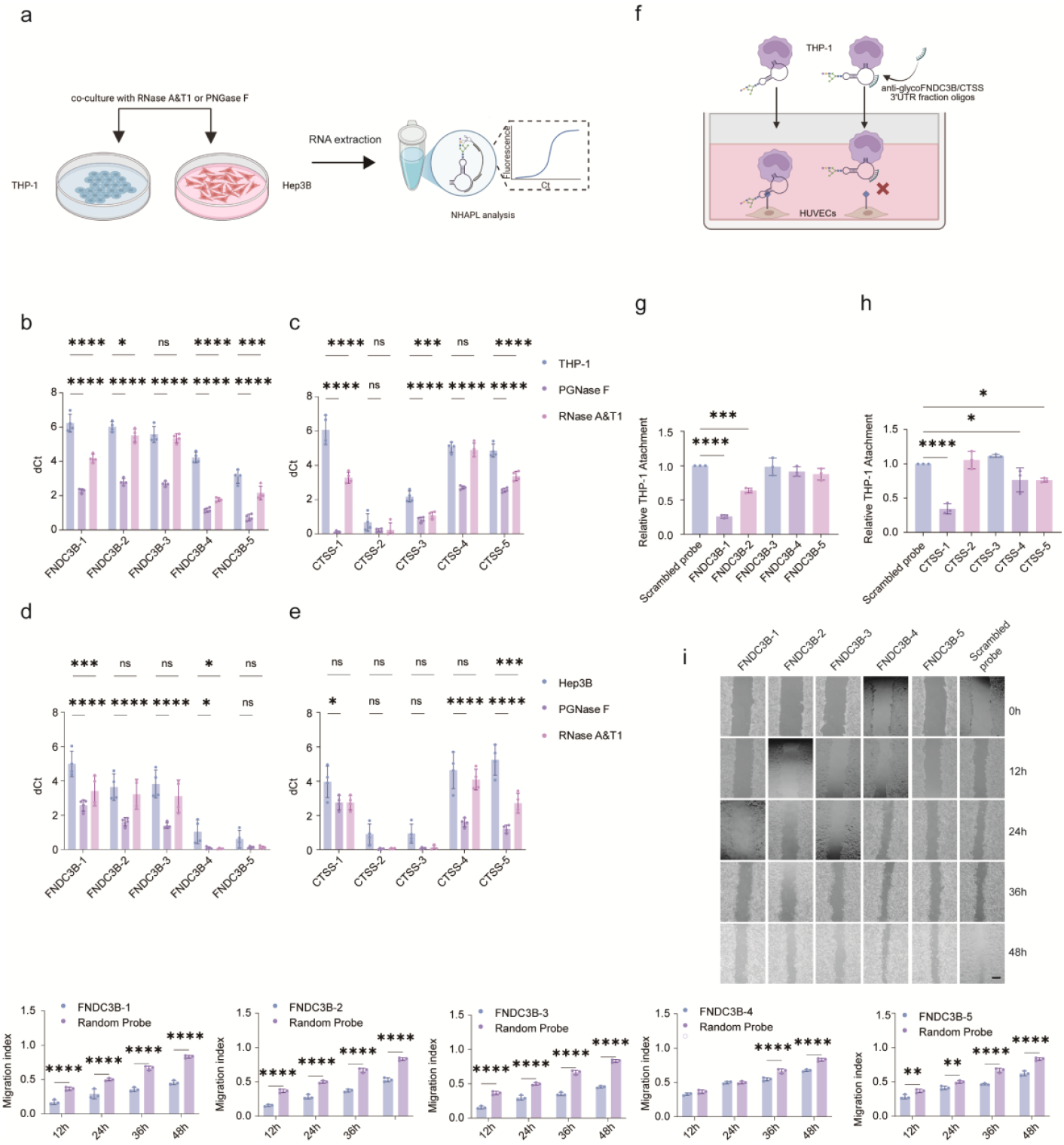
N-glycosylation modification of mRNA 3’UTR fractions. **a**, Schematic illustration showing that live THP-1 and Hep3B cells were treated with PNGase F or RNase to remove membrane glycans or RNA, respectively, followed by RNA extraction and NHAPL analysis. **b– e**, NHAPL analysis of glyco*FNDC3B* and glyco*CTSS* in THP-1 and Hep3B cells with or without enzymatic treatment. The left y-axis is the NHAPL signal dCt calculated by subtracting the Ct value of the RNA binding probes from that of RNA binding probes without RNA binding region (negative control, w/o RBR). (n = 4). **f**, Schematic showing that glyco*FNDC3B* and glyco*CTSS* localize on the cell surface and mediate THP-1 adhesion to vascular endothelial cells. **g, h**, Effects of individual antisense oligos complimentary to these membrane glyco3’UTR fragments on THP-1 adhesion compared with scrambled-oligo control. **i**, Wound-healing assay showing that cell surface glyco*FNDC3B* promotes Hep3B migration. Scale bar, 500 µm. Data shown as mean ± s.d. (n = 3). ns (not statistically significant), *(P < 0.05), ** (P < 0.01), *** (P < 0.001), **** (P < 0.0001).

Given the observed glyco3’UTR fragments correlation between THP-1 and Hep3B, we next investigated whether these glyco3’UTR fragments influence cellular functions. To perturb these glyco3’UTR fragments while avoiding interference with the protein encoded by the same gene’s mRNA, extracellular hybridization by antisense probes complementary to these cell surface glyco3’UTR fragments were employed. THP-1 cells were first fluorescent labeled, followed by incubating with each antisense probe set for 30 min, subsequently seeded onto confluent human umbilical vein endothelial cells (HUVECs) for adhesion (Fig. 5e). As shown in (Fig. 5f, g), after incubating THP-1 cells with these antisense oligos before co-culture with HUVECs, the adhesion ability of THP-s was significantly impaired, consistent with prior findings(Huang et al., 2020; N. Zhang et al., 2024). Compared with another work reporting that cell surface RNAs facilitate neutrophil recruitment to inflammatory sites in vivo by pretreating neutrophil with RNase to digest membrane RNA without selectivity(N. Zhang et al., 2024), we firstly validated that single target glycoRNA perturbance could interfere cell adhesion.

*FNDC3B* encodes a protein predominantly localized to the ER membrane and is implicated in migration and adhesion(Cai et al., 2012; Chen et al., 2010). It has also been linked to tumor progression in hepatocellular carcinoma(C. H. Lin et al., 2016; X. Y. Zhang et al., 2009). To further test the functional role of glyco*FNDC3B*, wound-healing assay were performed by live Hep3B cells co-cultured with five anti-*FNDC3B* oligonucleotides and a scrambled antisense oligo control, respectively. As shown in Fig. 5h, a marked reduction in migration was observed. These results indicate that cell surface glyco*FNDC3B* contributed in Hep3B migration.

### 4.5 Serum glycoRNA are potential disease markers

As glycoRNA can stably localize on the cell surface and resist RNase digestion (Fig. 2i, 5a-e) (T. Ren et al., 2025; N. Zhang et al., 2024), we next investigated whether they are also present in serum. As shown in Fig. 6a, 1 µl of serum was directly analyzed by multiplexed NHAPL without RNA extraction (see Methods for more information). To validate serum glycoRNA, we examined eight healthy young individuals (four males and four females, aged 15–25 years) and quantified five glycoRNA (U1, U3, U35, U8, and Y5, RNA binding probe without RNA binding region as negative control). The results revealed highly consistent expression among subjects, with a favorable Pearson correlation coefficient ranking from 0.87 to 1.0 (Fig. 6b, c). As shown in Extended Fig. 4 a, b, a twofold serial dilution of serum samples followed by multiplexed NHAPL analysis yielded a strong linear correlation, demonstrating that multiplexed NHAPL exhibit robust analytical ability in serum glycoRNA detection. Furthermore, serum glycoRNA remained stable after 10 freeze–thaw cycles (Fig. 6d) and incubation at 80°C for 10 min (Extended Fig. 5 a, b), indicating their robustness and reproducibility.

**Fig. 6.**
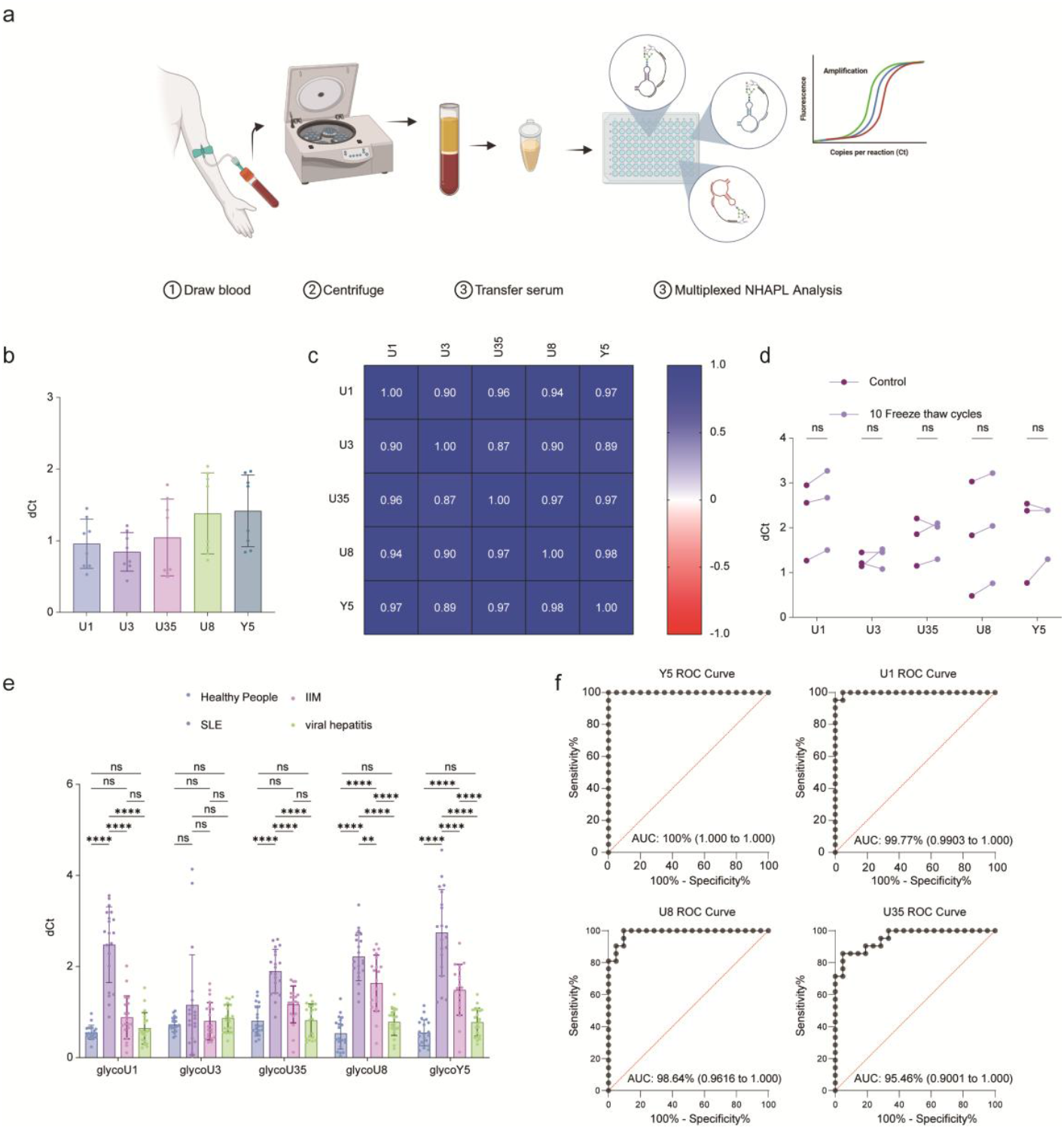
Serum glycoRNA as potential disease biomarkers. **a**, Workflow of multiplexed NHAPL analysis of serum glycoRNA without RNA extraction. **b**, Representative glycoRNA expression profiles from 8 healthy donors. **c**, Pearson correlation analysis showing high consistency of serum glycoRNA abundance among individuals. **d**, Stability test of serum glycoRNA after 10 freeze–thaw cycles. **e**, Comparison of 5 representative glycoRNA between serum from 20 healthy subjects, 20 SLE patients, 20 IIM and 20 viral hepatitis patients. The left y-axis is the NHAPL signal dCt calculated by subtracting the Ct value of the RNA binding probes from that of RNA binding probes without RNA binding region (negative control, w/o RBR). **f**, ROC curves for discrimination of SLE from healthy controls based on individual glycoRNA. Data shown as mean ± s.d. Each serum sample was analyzed with 2 technical replicates. ns (not statistically significant), *(P < 0.05), ** (P < 0.01), *** (P < 0.001), **** (P < 0.0001).

RNAs reported to undergo N-glycosylation are known to play critical roles in diverse cellular processes(Flynn et al., 2021). Among them, Y RNAs have attracted particular attention because of their well-established association with autoantigens in systemic lupus erythematosus (SLE)(Boccitto & Wolin, 2019; Marshak-Rothstein & Rifkin, 2007). To explore their diagnostic potential, we analyzed serum samples (n = 20 per group) from hospitalized patients with confirmed diagnoses of SLE, idiopathic inflammatory myopathies (IIM), viral hepatitis, as well as from healthy controls. Multiplexed NHAPL was applied to quantify glycoU1, glycoU3, glycoU35, glycoU8, and glycoY5, with an RNA-binding probe lacking the RNA-binding region serving as a negative control. Serum levels of glycoU1, glycoU35, glycoU8, and glycoY5 were markedly elevated in SLE patients compared with healthy donors and disease controls (Fig. 6e). Notably, glycoY5 achieved complete separation between SLE patients and healthy controls, and receiver operating characteristic (ROC) analysis demonstrated excellent diagnostic performance with an area under the curve (AUC) of 1.0 (Fig. 6f). Although serum glycoU8 and glycoY5 levels were also significantly increased in IIM patients relative to healthy controls, their levels in viral hepatitis patients remained comparable to those observed in healthy donors. This pattern indicates that elevated serum glycoRNA are not a general consequence of systemic inflammation or viral infection, but rather reflect autoimmune-associated immune dysregulation. Collectively, these findings position serum glycoRNA as biomarkers of autoimmune immune activation, with a preferential diagnostic relevance for SLE. Next, we examined the expression levels of these RNAs directly extracted from serum using conventional RT–qPCR. Given that no consensus exists regarding a stable endogenous reference RNA for serum-based RT–qPCR normalization, target RNA levels were normalized to the serum input volume (see Methods for details). Notably, no statistically significant differences were observed among SLE patients, IIM patients, viral hepatitis patients, and healthy donors using this conventional approach (Extended Fig. 6).

## 5 Discussion

The present work is the first one to analysis glycoRNA without sample restriction. We developed a homogenous detection method called NHAPL to analysis glycoRNA. NHAPL uses a sialic acid aptamer and an RNA-binding probe for dual recognition of the glycan and RNA portions of glycoRNA, respectively, followed by proximity ligation to generate intact lineage DNA product, which then serves as the template for qPCR amplification to transduce glycoRNA signal to Ct values. NHAPL exhibits several advantages: (1) the samples are easy to prepare, for example, native, unlabeled glycoRNA in various types of samples without metabolic labeling, cell fixing, oxidation. (2) the use of a DNA ligation for the detection reactions permits simultaneous analysis of large sets of glycoRNA by specifically design code sequences in the amplified segment for separate detection, for example by design RNA binding probes with different code primer site. (3) homogenous detection ability enables it a widely application in various samples, especially serum glycoRNA analysis for clinical diagnosis. (4) NHAPL exhibit high sensitivity by detection target glycoRNA from 160pg total RNA and 1µl serum, facilitating it a practical tool for glycoRNA study and methods development by further modification. (4) NHAPL amplifies glycoRNA detection signals through PCR, thereby achieving a dynamic range of 10 orders of magnitude (base 2). After validating the performance of NHAPL in total RNA, we further developed a multiplexed NHAPL for simultaneous analysis of a panel of glycoRNA. PLA has been applied for extracellular vesicles(B. Lin et al., 2021; Löf et al., 2016; Thulin et al., 2018) and virus(Gustafsdottir et al., 2006) analysis. Theoretically, NHAPL can also be used for glycoRNA in extracellular vesicles (T. Ren et al., 2025; Sharma et al., 2025) and virus (has not been reported yet).

Since glycoRNA have thus far been reported exclusively in the context of small non-coding RNAs(Flynn et al., 2021), we hypothesized and herein demonstrate for the first time that protein-coding RNAs are also subject to N-glycosylation. It has been reported that nuclear-encoded RNAs could present on the cell surface, namely membrane-associated extracellular RNAs (maxRNAs)(Huang et al., 2020). Inspired by this work, we applied NHAPL to investigate whether these maxRNAs are N-glycosylation modified. Unexpectedly, we discovered that 3’ untranslated region (3’UTR) fragments from the protein-coding RNAs FNDC3B and CTSS exist as glycosylated RNAs on the cell surface, thereby providing the first demonstration that protein-coding RNAs, rather than only small non-coding RNAs, can undergo N-glycosylation. Functionally, these glycoRNA modulate monocyte adhesion to vascular endothelial cells, extending recent findings linking surface glycoRNA to P-selectin–mediated cell adhesion(N. Zhang et al., 2024). Here, we have firstly extended glycoRNA research from general study to single molecule function investigation. What’s more, we further uncover that glyco*FNDC3B* also contribute hepatoma cell migration like its coding protein(C. H. Lin et al., 2016). Our findings not only establish mRNA-based substate for modification with sialic acid-containing N-linked glycans but also raise the interesting possible relationship between the glyco*FNDC3B*/*CTSS* RNA and *FNDC3B, CTSS* protein and even other mRNA undiscovered yet. It is interesting to consider why mRNA N-glycosylation had not been found previously(Flynn et al., 2021). This is probably due to a poly(A) enrichment strategy missed fractionated mRNA(Flynn et al., 2021). Given that the full composition of membrane-associated RNAs is not yet clearly defined, the findings derived from antisense oligonucleotide blockade of surface glycoRNA may not fully capture glycoRNA-specific effects. It is possible that the observed phenomena partially result from interference with other membrane RNAs independent of glycosylation.

Previous work has reported that cell-free RNA (cfRNA) is highly fragmented, containing no detectable 18S and 28S rRNA peaks(Nesselbush et al., 2025). The median concentration of cfRNA was 220 pg per millilitre plasma(Nesselbush et al., 2025), corresponding to the optimal total RNA input of 160pg (Fig. 1b), in which small RNA with N-glycosylation accounts for a very small proportion, rendering glycoRNA detection in serum without RNA extraction feasible with NHAPL. Here, we applied multiplexed NHAPL to validate the stable presence of serum glycoRNA and reproducibly consistent expression level among individuals. Next, we uncovered disease-associated glycoRNA signatures in systemic lupus erythematosus (SLE). Serum glycoY5 and glycoU1 exhibited exceptional diagnostic performance, achieving near-complete discrimination between SLE patients and healthy controls. In contrast, by incorporating autoimmune (SLE and IIM) and non-autoimmune (viral hepatitis) cohorts, we show that these glycoRNAs are preferentially enriched in SLE rather than broadly elevated in inflammatory states. Importantly, conventional RT-qPCR analysis of total serum RNA failed to detect corresponding differences across disease groups, underscoring that glycoRNA modification, rather than transcript abundance, represents a distinct and previously underappreciated layer of disease-associated regulation. A recent research found heterogeneity in serum-isolated glycoRNA signals in a small set of healthy controls and patients with SLE using rPAL, but the overall changes are not statistically significant(Graziano et al., 2025). This discrepancy may stem from differences in both sample origin and detection methodology. The earlier study recruited patients with suspected neuroinflammatory disorders(Graziano et al., 2025), whereas our samples were derived from clinically confirmed SLE patients. Methodologically, the rPAL assay employed in the previous work remains a blotting-based approach akin to RNA immunoblotting. However, NHAPL employs a PCR-based signal amplification strategy, exhibiting superior stability and sensitivity compared with blotting-based methods, which may underlie the divergent detection outcomes.

U1 small nuclear ribonucleoprotein (U1 snRNP) is a key autoantigen in systemic lupus erythematosus (SLE). Autoantibodies against U1, mainly anti-Sm and anti-RNP, serve as important diagnostic markers(Migliorini, Baldini, Rocchi, & Bombardieri, 2005). The Ro particle, a frequent autoimmune target in systemic lupus erythematosus (SLE), contains 52- and 60-kD proteins bound to Y1–Y5 RNAs, among which Y5 has been increasingly implicated in autoantibody responses, highlighting its potential role in SLE pathogenesis(Benchetrit, Gandy, Tan, & Sullivan, 1989). Here, we firstly report that these SLE associated RNAs, especially glycoY5 and U1, presented an excellent sensitivity SLE diagnosis (AUC: 100% for glycoY5, 99.77% for glycoU1) compared with traditional auto-antibody biomarkers, for example, anti-sm antibodies are detectable in 5-30% SLE patients, anti-RNP are detectable in 25–47% of SLE patients(Migliorini et al., 2005). Notably, miR155 has been validated to be N-glycosylation modified (Fig. 2b), which is a chronic infections of hepatitis B or C viruses biomarker(Alizadeh et al., 2015; Kawaguchi et al., 2016; Mohammadian, Pilehvar-Soltanahmadi, Zarghami, Akbarzadeh, & Zarghami, 2017). Using SPCgRNA platform, a penal of miRNAs associated with pancreatic cancers have been identified as glycoRNA(J. J. Li et al., 2023). While the present study focused on cross-sectional cohorts, future studies incorporating pre- and post-treatment samples may clarify whether dynamic changes in serum glycoRNA reflect disease activity or treatment efficacy in SLE. Taken above, glycoRNA have a promising application as clinical diagnosis.

The present version of NHAPL platform is intrinsically constrained by its reliance on sequence-specific RNA hybridization and sialic acid–binding aptamers, limiting detection to predefined, sialylated glycoRNAs. Consequently, glycoRNAs bearing other terminal glycan structures, such as fucosylation or non-sialylated N-glycans, are not captured by the present platform. This constraint reflects a deliberate design choice prioritizing analytical specificity and assay simplicity, rather than a fundamental limitation of the proximity ligation strategy. Future expansion of the method using alternative or multiplexed glycan-binding aptamers may enable broader glycoform coverage and further enhance the versatility of NHAPL.

In summary, this study introduces NHAPL as a versatile and sensitive platform for glycoRNA detection and provides compelling evidence that mRNA 3′UTRs can undergo N-glycosylation with functional consequences. By linking glycoRNA biology to autoimmune disease, particularly SLE, our findings not only expand the conceptual framework of RNA glycosylation but also highlight the translational potential of glycoRNAs as disease-specific liquid biopsy biomarkers.

## Data Availability

All data produced in the present study are available upon reasonable request to the authors

## 7 Acknowledgments

## Author contributions

Conceptualization: Jie Gui. Methodology: Jie Gui. Investigation: Jie Gui, Meng Zhang, Ziwei Kan, Xiaojuan He, Meipei Gao, Jian Han, Qiongfang Wang, Shengyao Zhang, Junyi Hu, Wenyi Qin, Boyue Huang, Zhongjun Wu, Jianhua Ran. Visualization: Jie Gui. Supervision: Jianhua Ran. Writing—original draft: Jie Gui. Writing— review & editing: Jie Gui, Meng Zhang, Ziwei Kan, Jian Han, Boyue Huang, Zhongjun Wu, Jianhua Ran. Funding: Xiaojuan He, Meipei Gao, Boyue Huang, Zhongjun Wu, Jianhua Ran.

## Data and materials availability

All data are available in the main text or the supplementary materials.

