## Supplementary material for "NHAPL analysis of glycoRNA reveals sialic acid-containing glycosylated mRNA 3′UTRs and enables sensitive SLE diagnostics": Supplementary data.pdf

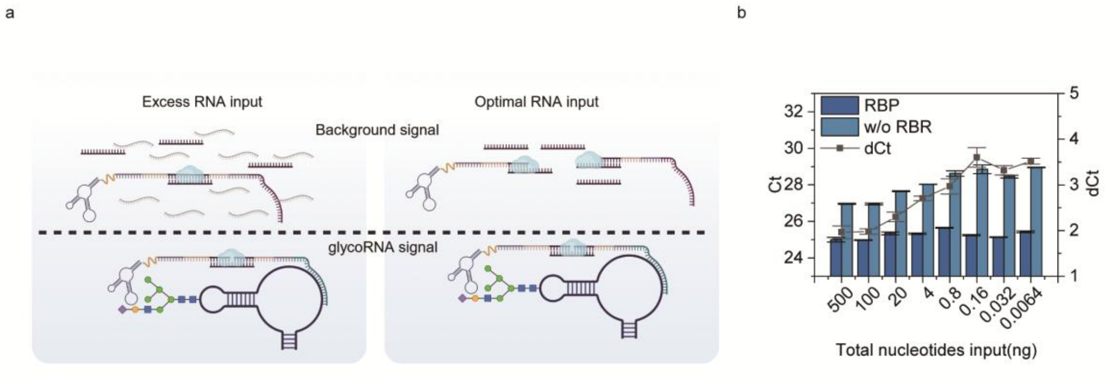


**Extended Fig. 1 a,** Diagram showing that excessive total RNA increases background by hindering connector binding, while optimal input minimizes this effect. **b,** NHAPL Ct and dCt values across varying salmon sperm DNA inputs with a fixed RNA input at 160pg. For less than 160pg RNA input, no salmon sperm DNA was added; The left y-axis is the NHAPL signal Ct. The right y-axis is the NHAPL signal dCt calculated by subtracting the Ct value of the RNA binding probes for U1 (RBP) from that of RNA binding probes without RNA binding region (negative control, w/o RBR). Data shown as mean ± s.d. (n = 3).

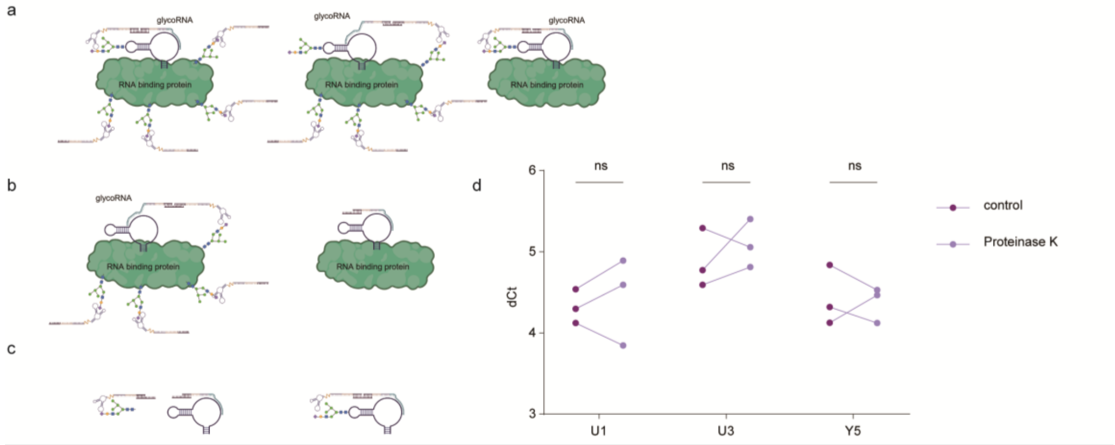


**Extended Fig. 2 a, b** Diagram showing the impact of RNA binding protein on NHAPL analysis for glycoRNA. **c,** Illustration showing the impact of free glycan on NHAPL analysis for glycoRNA. **d**, NHAPL analysis for cellular RNA sample with or without proteinase K treatment. The left y-axis is the NHAPL signal dCt calculated by subtracting the Ct value of the RNA binding probes (glycoU1, U3 and Y5) from that of RNA binding probes without RNA binding region (negative control, w/o RBR). Data shown as mean ± s.d. (n = 3). ns (not statistically significant).

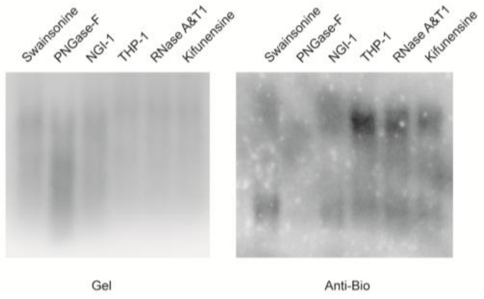

**Extended Fig. 3,** Total RNA expression level after pharmacological and enzymatic methods to remove the glycan or RNA moiety by metabolic labeling with Ac4ManNAz followed by biotinylation and RNA blotting.

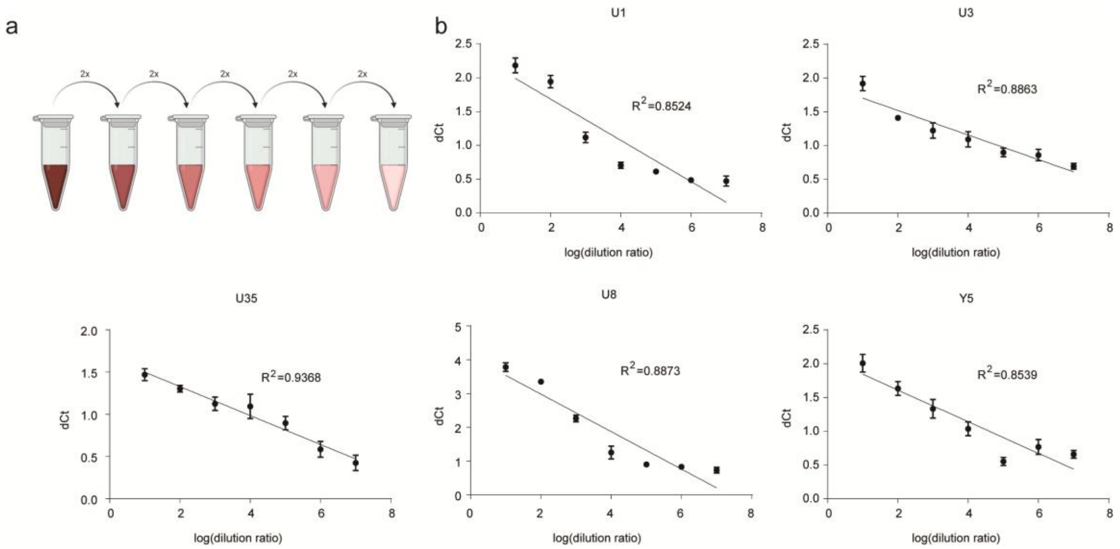


**Extended Fig. 4 a,** Diagram illustrating a twofold serial dilution of serum samples followed by multiplexed NHAPL analysis. **b,** Multiplexed NHAPL dCt values across a serum dilution series; The left y-axis is the NHAPL signal dCt calculated by subtracting the Ct value of the RNA binding probes for corresponding glycoRNA from that of RNA binding probes without RNA binding region (negative control). Considering the qPCR signals were logarithmic in nature, the serum sample concentration was log-transformed and plotted on the x-axis. Data shown as mean ± s.d. (n = 3).

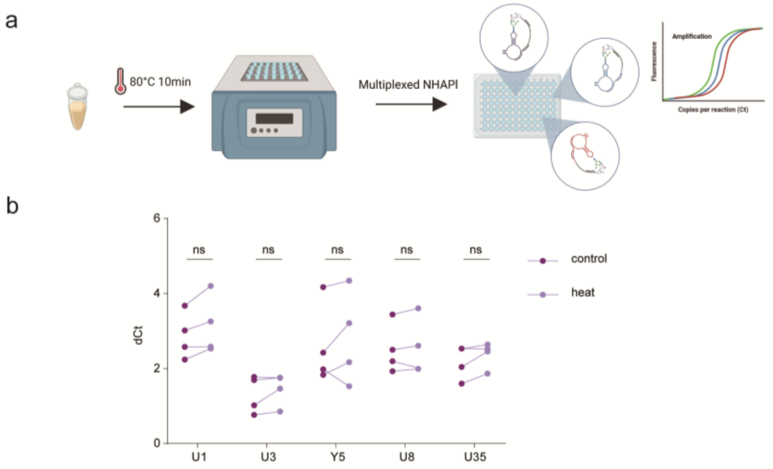


**Extended Fig. 5 a,** Schematic illustration showing that serum was incubated at 80°C for 10 min, followed by multiplexed NHAPL analysis with untreated serum as control. **b,** Evaluation of the effect of serum heat treatment on multiplexed NHAPL dCt values; The left y-axis is the NHAPL signal dCt calculated by subtracting the Ct value of the RNA binding probes for corresponding glycoRNA from that of RNA binding probes without RNA binding region (negative control, w/o RBR). Data shown as mean ± s.d. (n = 3). ns (not statistically significant).

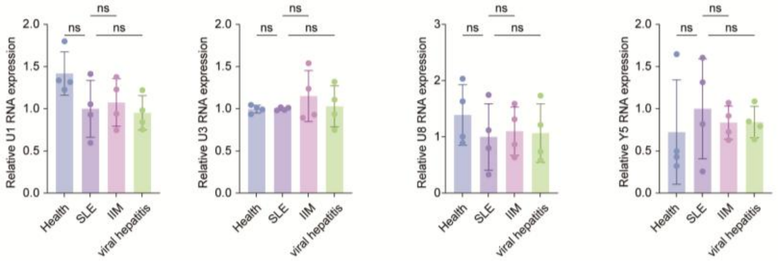


**Extended Fig. 6** RT-qPCR results of U1, U3, U8 and Y5 RNA for serum extracted total RNA. The expression levels of target RNA were directly normalized to serum input in RNA extraction procedures. Data shown as mean ± s.d. (n = 4). ns (not statistically significant).

| Table S1. anti-sense oligos used in this work | |
|---|---|
| Name | Sequence (5' -> 3') |
| anti-CTSS-1 | ACAAAAGGCTATTAGGGGTC |
| anti-CTSS-2 | TGGTATGTGGCAATGATAGA |
| anti-CTSS-3 | TTATTAATAGTTGCCTAGGA |
| anti-CTSS-4 | AAAGTTGCTAAACCCCATCA |
| anti-CTSS-5 | ATCTGAGTCCTTAGGACATT |
| anti-FNDC3B-1 | ACTACAGATGTTGCCAAGGA |
| anti-FNDC3B-2 | ATCTCAGCATTTTCTTCCAA |
| anti-FNDC3B-3 | AAATGCAGATGCTGTGGTCA |
| anti-FNDC3B-4 | TCAGCTTTGTACTTCACACA |
| anti-FNDC3B-5 | AGTTTGCTTCACATGACTGG |

| Table S2. Proximity probes, connector and primers used in this work | |
|---|---|
| Name | Sequence (5' -> 3') |
| connector | AAAGAATGATGACCCTCTTGCTAAAA |
| glycoCTSS-1 probe | 5phos/GTCATCATTCGAATCGTACTGCAATCGGGTATT**AAAAAAAAAA**ACAAAAGGCTATTAGGGGTC |
| glycoCTSS-2 probe | 5phos/GTCATCATTCGAATCGTACTGCAATCGGGTATT**AAAAAAAAAA**TGGTATGTGGCAATGATAGA |
| glycoCTSS-3 probe | 5phos/GTCATCATTCGAATCGTACTGCAATCGGGTATT**AAAAAAAAAA**TTATTAATAGTTGCCTAGGA |
| glycoCTSS-4 probe | 5phos/GTCATCATTCGAATCGTACTGCAATCGGGTATT**AAAAAAAAAA**AAAGTTGCTAAACCCCATCA |
| glycoCTSS-5 probe | 5phos/GTCATCATTCGAATCGTACTGCAATCGGGTATT**AAAAAAAAAA**ATCTGAGTCCTTAGGACATT |
| glycoFNDC3B-1 probe | 5phos/GTCATCATTCGAATCGTACTGCAATCGGGTATT**AAAAAAAAAA**ACTACAGATGTTGCCAAGGA |
| glycoFNDC3B-2 probe | 5phos/GTCATCATTCGAATCGTACTGCAATCGGGTATT**AAAAAAAAAA**TCTCAGCATTTTCTTCCAA |
| glycoFNDC3B-3 probe | 5phos/GTCATCATTCGAATCGTACTGCAATCGGGTATT**AAAAAAAAAA**AAATGCAGATGCTGTGGTCA |
| glycoFNDC3B-4 probe | 5phos/GTCATCATTCGAATCGTACTGCAATCGGGTATT**AAAAAAAAAA**TCAGCTTTGTACTTCACACA |
| glycoFNDC3B-5 probe | 5phos/GTCATCATTCGAATCGTACTGCAATCGGGTATT**AAAAAAAAAA**AGTTTGCTTCACATGACTGG |
| Glycan probe | TAGGGAATTCGTCGACGGATCCCGTGGCGTCTGCAACGGAAAAGAATTTATCTTGTCCTGCAGGTCGACGCATGCGCCG**AAAAAAAAAA**GTGACTTCGTGGAACTATCTAGCGTGTAGTGAGTGGGCATGTAGCAAGAGG |

| | |
|---|---|
| Glycan probe using DNA with scrambled sequence | CGGCGCATGCGTCGACCTGCAGGACAAGATAAATTCTTTTCCGTTGCAGACGCCACGGGATCCGTCGACGAATTCCCTA**AAAAAAAAAA**GTGACTTCGTGGAACTATCTAGCGTGTAGTGAGTGGGCATGTAGCAAGAGG |
| Glycan probe without aptamer (w/o G) | **AAAAAAAAAA**GTGACTTCGTGGAACTATCTAGCGTGTAGTGAGTGGGCATGTAGCAAGAGG |
| glycoU1 probe | 5phos/GTCATCATTCGAATCGTACTGCAATCGGGTATT**AAAAAAAAAA**CTGGGAAAACCACCTTCGTGATCATGGTATCTCCCCTGCCAGGTAAGTAT |
| glycoU3 probe | 5phos/GTCATCATTCGAATCGTACTGCAATCGGGTATT**AAAAAAAAAA**CTCCCCAATACGGAGAGAAG |
| glycoU35 probe | 5phos/GTCATCATTCGAATCGTACTGCAATCGGGTATT**AAAAAAAAAA**AGACCATCGTGAGATAAG |
| glycoY5 probe | 5phos/GTCATCATTCGAATCGTACTGCAATCGGGTATT**AAAAAAAAAA**GGGAGACAATGTTAAATC |
| Multiplexed U1 probe | 5phos/GTCATCATTCCGTGACAGTGGCAGATATAAC**AAAAAAAAAA**CTGGGAAAACCACCTTCGTGATCATGGTATCTCCCCTGCCAGGTAAGTAT |
| Multiplexed U1 Primer | GTTATATCTGCCACTGTCACG |
| Multiplexed U3 probe | 5phos/GTCATCATTCCGTGAACCGTTATTTGGGTAC**AAAAAAAAAA**CTCCCCAATACGGAGAGAAG |
| Multiplexed U3 Primer | GTACCCAAATAACGGTTCACG |
| Multiplexed U35a probe | 5phos/GTCATCATTCCGCAGGCAGATCGACCTAGTT**AAAAAAAAAA**AGACCATCGTGAGATAAG |
| Multiplexed U35a Primer | AACTAGGTCGATCTGCCTGCG |
| Multiplexed U8 probe | 5phos/GTCATCATTCCGCGAGCGTACTATACATAAC**AAAAAAAAAA**GTTCTAATCTGCCCTCCGGAGGAGGAACAGGTAAGGATTATCCCACC |
| Multiplexed U8 Primer | GTTATGTATAGTACGCTCGCG |
| Multiplexed Y5 probe | 5phos/GTCATCATTCCGTGCTGCGAGAGTATTATCT**AAAAAAAAAA**GGGAGACAATGTTAAATC |
| Multiplexed Y5 Primer | AGATAATACTCTCGCAGCACG |
| Multiplexed-w/o R probe | 5phos/GTCATCATTCCGAACTATGCTGACAGTACCG**AAAAAAAAAA** |
| Multiplexed-w/o R-primer | CGGTACTGTCAGCATAGTTCG |
| Primer-F (Universal primer) | GTGACTTCGTGGAACTATCTAGCG |
| Primer-R (single NHAPL) | AATACCCGATTGCAGTACGATTC |
| RNA binding probe without hydridazation (w/o R) | 5phos/GTCATCATTCGAATCGTACTGCAATCGGGTATT**AAAAAAAAAA** |
| glycomi155 probe | 5phos/GTCATCATTCGAATCGTACTGCAATCGGGTATT**AAAAAAAAAA**ACCCCTATCACGATTAGCATTAA |

Note: Aptamer and RNA binding regions are underlined. Poly(A) linker is bolded. glycoRNA specific code in RNA binding probes for multiplexed NHAPL are underlined with a dot line.

| Table S3. RT-qPCR primer sequences | |
|---|---|
| Name | Sequence (5' -> 3') |
| Y5 primer-F | agttggtccgagtgttgt |
| Y5 primer-R | aaaacagcaagctagtcaagc |
| U8-F | atcgtcaggtgggataatcctt |
| U8-R | gagacgttaatcacgtttcatgc |
| U3-F | actttcagggatcatttctatagtgtg |

| | |
|---|---|
| U3-R | gaaagccggcttcacgct |
| U1-F | atacttacctggcaggggagata |
| U1-R | caggggaaagcgcgaac |

| Table S4. Demographic characteristics of serum donors | | | |
|---|---|---|---|
| Series number | Gender | Age range (years) | Gourp |
| 1 | Male | 11~15 | healthy donors |
| 2 | Male | 11~15 | healthy donors |
| 3 | Female | 11~15 | healthy donors |
| 4 | Male | 11~15 | healthy donors |
| 5 | Female | 11~15 | healthy donors |
| 6 | Male | 16~20 | healthy donors |
| 7 | Male | 16~20 | healthy donors |
| 8 | Female | 21~25 | healthy donors |
| 9 | Female | 21~25 | healthy donors |
| 10 | Male | 21~25 | healthy donors |
| 11 | Female | 26~30 | healthy donors |
| 12 | Female | 26~30 | healthy donors |
| 13 | Male | 31~35 | healthy donors |
| 14 | Male | 31~35 | healthy donors |
| 15 | Female | 36~40 | healthy donors |
| 16 | Female | 36~40 | healthy donors |
| 17 | Male | 41~45 | healthy donors |
| 18 | Female | 41~45 | healthy donors |
| 19 | Female | 41~45 | healthy donors |
| 20 | Male | 41~45 | healthy donors |
| 1 | Female | 30~31 | SLE patients |
| 2 | Female | 30~31 | SLE patients |
| 3 | Female | 30~31 | SLE patients |
| 4 | Female | 36~40 | SLE patients |
| 5 | Female | 36~40 | SLE patients |
| 6 | Female | 36~40 | SLE patients |
| 7 | Female | 36~40 | SLE patients |
| 8 | Female | 41~45 | SLE patients |
| 9 | Female | 41~45 | SLE patients |
| 10 | Female | 46~50 | SLE patients |
| 11 | Female | 46~50 | SLE patients |
| 12 | Female | 46~50 | SLE patients |
| 13 | Female | 46~50 | SLE patients |
| 14 | Female | 51~55 | SLE patients |
| 15 | Female | 51~55 | SLE patients |
| 16 | Female | 51~55 | SLE patients |
| 17 | Female | 56~60 | SLE patients |
| 18 | Female | 61~65 | SLE patients |
| 19 | Female | 61~65 | SLE patients |
| 20 | Female | 61~65 | SLE patients |
| 1 | Male | 31~35 | IIM |

| | | | |
|---|---|---|---|
| 2 | Female | 51~55 | IIM |
| 3 | Male | 46~50 | IIM |
| 4 | Male | 31~35 | IIM |
| 5 | Female | 31~35 | IIM |
| 6 | Female | 56~60 | IIM |
| 7 | Male | 60~65 | IIM |
| 8 | Female | 31~35 | IIM |
| 9 | Male | 41~45 | IIM |
| 10 | Female | 46~50 | IIM |
| 11 | Female | 31~35 | IIM |
| 12 | Male | 36~40 | IIM |
| 13 | Female | 36~40 | IIM |
| 14 | Female | 46~50 | IIM |
| 15 | Female | 36~40 | IIM |
| 16 | Male | 36~40 | IIM |
| 17 | Male | 31~35 | IIM |
| 18 | Female | 36~40 | IIM |
| 19 | Female | 41~45 | IIM |
| 20 | Male | 46~50 | IIM |
| 1 | Male | 31~35 | viral hepatitis |
| 2 | Male | 36~40 | viral hepatitis |
| 3 | Female | 51~55 | viral hepatitis |
| 4 | Male | 36~40 | viral hepatitis |
| 5 | Male | 41~45 | viral hepatitis |
| 6 | Female | 31~35 | viral hepatitis |
| 7 | Male | 46~50 | viral hepatitis |
| 8 | Female | 51~55 | viral hepatitis |
| 9 | Female | 51~55 | viral hepatitis |
| 10 | Female | 41~45 | viral hepatitis |
| 11 | Male | 56~60 | viral hepatitis |
| 12 | Female | 36~40 | viral hepatitis |
| 13 | Male | 31~35 | viral hepatitis |
| 14 | Male | 46~50 | viral hepatitis |
| 15 | Female | 31~35 | viral hepatitis |
| 16 | Female | 56~60 | viral hepatitis |
| 17 | Male | 41~45 | viral hepatitis |
| 18 | Female | 56~60 | viral hepatitis |
| 19 | Male | 26~30 | viral hepatitis |
| 20 | Male | 36~40 | viral hepatitis |

Note: The 20 serum samples from hospitalized patients were confirmed with diagnosis of SLE.
